# Prospective metrological validation of multifrequency bioelectrical impedance analysis against volumetric imaging to identify sarcopenia in head and neck cancer patients undergoing radiation therapy

**DOI:** 10.1101/19006668

**Authors:** Aaron J. Grossberg, Crosby D. Rock, Jared Edwards, Abdallah S.R. Mohamed, Debra Ruzensky, Angela Currie, Patricia Rosemond, Jack Phan, G. Brandon Gunn, Steven J. Frank, William H. Morrison, Adam S. Garden, Clifton D. Fuller, David I. Rosenthal

## Abstract

**Importance:** Depleted skeletal muscle mass (sarcopenia) is associated with decreased survival and cancer control in head and neck cancer patients treated with radiotherapy. There is a need for validated measures of body composition that can be implemented in routine clinical workflow.

**Objective:** To validate the use of bioelectrical impedance analysis (BIA) for body composition analysis and diagnosis of sarcopenia in head and neck cancer patients.

**Design:** In this prospective observational cohort study, baseline 50 patients with head and neck cancer undergoing radiation therapy (RT) were enrolled between February 2016 and March 2017. Baseline BIA measures of skeletal muscle (SM) mass, fat-free mass (FMM), and fat mass (FM) were compared to CT-based estimates of body composition using linear regression. Sex-specific BIA-derived thresholds for sarcopenia were defined by the maximum Youden Index on receiver operator characteristic (ROC) curves of BIA against CT-defined sarcopenia. Changes in body composition across treatment were compared against changes in body weight using linear regression.

**Participants:** In total, 50 patients with pathologically confirmed stage I to IVB non-metastatic head and neck cancer treated with definitive radiation therapy were enrolled.

**Setting:** Single academic referral center.

**Main Outcome and Measure:** The primary outcome was relative agreement between baseline lean body mass and fat body mass predicted from BIA measurement and CT imaging.

**Results:** Of the 48 evaluable patients 16 (33.3%) were sarcopenic at baseline based on CT analysis. BIA measures of body composition were strongly correlated with CT measures: SM mass (*r* = 0.97; R^2^ = 0.94; *p* < 0.0001), FFM (*r* = 0.97; R^2^ = 0.94; *p* < 0.0001) and FM (*r* = 0.95; R^2^ = 0.90; *p* < 0.0001). Relationship with normalized indices of SM mass, FFM, and FM was similar between BIA and CT, but not BIA and body mass index (BMI). Patients lost a mean of 5.7 ± 5.8 kg during treatment, of which 1.5 ± 1.9 kg was SM, 2.6 ± 3.3 kg was FFM, and 2.2 ± 2.6 kg was FM. Eight additional patients developed sarcopenia by the end of RT.

**Conclusions:** BIA provides accurate estimates of body composition in head and neck cancer patients. Implementation of BIA in clinical practice may identify patients with sarcopenia.

**Trial Registration:** ClinicalTrials.gov identifier: NCT02615275

## Background

Head and neck cancer patients with decreased muscle mass, or sarcopenia, exhibit poor survival and locoregional cancer control as well as impaired tolerance of RT and increased chemotherapy toxicity.^1-6^ Weight loss itself shows little correlation with oncologic or functional outcomes, and weight-derived metrics, such as body mass index (BMI), are unable to differentiate between the lean and adipose contributions to body mass.^7^ The gold standard for body composition measurement is dual energy x-ray absorptiometry (DEXA), with more recent data validating the use of magnetic resonance imaging (MRI), or CT.^8-10^ However, these modalities are expensive, involve radiation exposure (in the case of CT and DEXA), and are often not pragmatic or available at radiation treatment centers. Furthermore, body composition analysis of CT or MRI scans is not routinely performed, so this information is unavailable for clinical use. Therefore, there is a need for body composition evaluation that can be rapidly implemented into routine clinical workflow.

BIA offers a non-invasive, cost-effective method for serially measuring body composition.^11^ BIA measures impedance to a weak electric current to estimate the relative fat and water content of the body. In a healthy patient population, BIA estimates of body composition did not differ significantly from estimates derived from the four-compartment model, as shown by high coefficients of determination (R^2^, 0.94-0.98) and low root mean square error (RMSE, −0.2-0.3 kg) across multiple ethnic populations.^12^ Head and neck cancer patients represent a more challenging population given wide fluctuations in hydration status, nutrition, and muscle mass. The accuracy of BIA in a head and neck cancer population has not been previously evaluated.

The primary objective of this study was to validate eight-electrode multifrequency BIA as a means to assess body composition against CT-based estimates in a head and neck cancer population undergoing RT. CT-based estimates were selected as the gold standard due to the previously described association between this measure of body composition and survival in this patient population. Secondary objectives were to serially examine the change in weight and body composition during RT using BIA.

## Materials and methods

### Study Design

Between February 2016 and March 2017, 50 patients were enrolled at The University of Texas MD Anderson Cancer Center during their treatment planning visits. Patients eligible for enrollment were age ≥ 18 years with pathologically confirmed head and neck cancer, AJCC 7^th^ edition stage Tx-T4, N0-3, M0, dispositioned to RT to a dose of ≥ 60 Gy either as single modality or as part of a multimodality treatment approach. Additionally, they required a staging PET/CT scan within a 30-day period prior to initiation of RT. Exclusion criteria included prior overlapping RT, pregnancy, or concomitant medical conditions known to cause cachexia or sarcopenia. All patients underwent treatment per consensus recommendation by multidisciplinary tumor board, without regard for study inclusion. This study was approved by the institutional review board of the University of Texas MD Anderson Cancer Center. Written informed consent was obtained from all study participants.

Serial BIA measures were performed at the pre-treatment quality assurance visit and during weekly on-treatment clinic visits. All patients undergoing RT received institutional standard pre-treatment speech and swallow evaluations and weekly dietary counseling by licensed dietitians. Feeding tubes were placed only therapeutically; no prophylactic feeding tubes were offered. Patient demographics and treatment and tumor characteristics were abstracted from the electronic medical record.

### Bioelectrical Impedance Analysis

Body composition was measured using the FDA-cleared SECA medical Body Composition Analyzer (mBCA) 515 scale (https://www.seca.com/en_no/products/all-products/product-details/seca515.html; seca gmbh & co. kg, Hamburg, Germany). One of three possible hand positions was selected such that the angle between the body and arms was as close to 30 degrees as possible. Impedance is measured using a 100 μA current delivered at 19 frequencies ranging from 1-1000 kHz.^12^ To avoid additional barriers to adequate nutrition and exercise, participants were not instructed to alter their food intake or activity prior to measurement. For each measure, patients removed their shoes and socks and emptied their pockets. Estimations of body composition were based on equations previously validated in healthy adults and incorporated into vendor-supplied SECA analytics 115 software (SECA North America, Chino, CA).^12^ BIA data were measured prior to RT initiation (baseline), then at weekly clinic visits during treatment. Measurements were stored outside the patients’ medical records and were not used for medical decision making.

### CT Image Analysis

Cross-sectional images of the third lumbar vertebrae (L3) were extracted from the CT component of pre-RT whole-body diagnostic PET-CT scans for each participant.^9,13-15^ Both SM mass and FM were calculated from L3 contours, as previously described.^9^ The mean cross-sectional area of muscle and adipose tissue was divided by the square of height in meters to normalize for patient height and reported as the lumbar SM index (SMI) or adipose index (ADI).^9,13,15^ SM depletion was defined *a priori* as an SMI of less than 52.4 for men and less than 38.5 for women.^1,15,16^

### Statistical analysis

Data are presented as means ± SD. Differences between groups were assessed using the 2-tailed Student’s *t* test (for continuous variables) and Pearson χ2 test or Fisher’s Exact Test (for categorical variables). Statistical significance was set at *p* < 0.05.

For the primary outcome, we used linear regression analysis to determine the relative agreement between baseline lean body mass and fat mass predicted from impedance measurement and CT imaging. The Pearson correlation, *r*, between BIA and CT imaging estimates of lean and fat body mass before RT as well as the coefficient of determination, R^2^, were calculated. Both sexes were included in correlation calculations as these relationships are age and sex invariant.^8^ RMSE analysis was utilized to quantify the average error from BIA-derived body composition calculations. The following BIA-derived body composition estimates were compared against CT-measured tissue Cross-Sectional Area (CSA): SM mass, FFM, and FM. Comparisons of SM mass against CT CSA were repeated for each body segment (torso, R & L legs, R & L arms). Height-normalized BIA estimates were compared against CT-derived SMI and ADI as well as BMI. To identify BIA-based thresholds for sarcopenia, ROC analyses were performed using the sex-specific CT-based sarcopenia definitions as ground truth. BIA sarcopenia thresholds were defined by identifying the optimal threshold from the ROC analyses based on maximum Youden Index—the value of the tested variable at which the sum of sensitivity and specificity is greatest. To evaluate whether weight loss during RT reflects changes in body composition, the changes in each BIA-measured tissue compartment and BMI were calculated by subtracting the baseline value from the measurement during the final week of treatment. R^2^ and RMSE were calculated from linear regression analysis comparing change in SM mass, FFM, and FM to change in BIA.

## Results

### Patient Characteristics

Fifty patients were enrolled, and 48 patients remained in the study and evaluable. Two patients withdrew consent to the study, and no data were collected. Two other patients requested to withdraw from the study because their treatment occurred offsite; their baseline data were included in analysis. The study population characteristics were comparable with the overall population of head and neck cancer patients treated with RT. Patients had a mean age of 60.2 (12.2) years at enrollment. The patient sample was predominately male (40 men [83.3%] vs 8 women [16.7%]) with mean BMI of 30.0 (4.5) for men and 24.3 (5.4) for women. The oropharynx was the most common primary cancer site, with 26 cases (54.2%). The majority of patients (28 [58.3%]) had multiple involved lymph nodes at diagnosis (AJCC 7^th^ edition N2-3). Human papillomavirus was detected by p16 immunohistochemistry or HPV polymerase chain reaction in all 26 oropharynx cancer patients (54.2%) and was not tested in 19 other patients (39.6%). Forty-seven of 48 patients (97.9%) had ECOG PS 0-1. Patient and treatment characteristics are outlined in Table 1.

**Table 1.** Patient and Treatment Characteristics. Separated by sex. Abbreviations: BMI, body mass index; SD, standard deviation; CUP, cancer of unknown primary; HPV, human papillomavirus; ECOG PS, Eastern Cooperative Oncology Group Performance Status; SM, skeletal muscle. P-values calculated using Student’s t-test for continuous variables, χ2 or Fisher’s Exact Test for categorical variables.

|  | Men<br>(n= 40, 83.3%) |  | Women<br>(n= 8, 16.7%) |  |  |
| --- | --- | --- | --- | --- | --- |
|  | mean | SD | mean | SD | P |
| Age (y) | 60.0 | 10.7 | 52.1 | 16.7 | 0.23 |
| Height (m) | 1.79 | 0.06 | 1.61 | 0.06 | <.0001 |
| Weight (kg) | 96.6 | 16.6 | 63.2 | 11.7 | <.0001 |
| BMI (kg/m2) | 30.0 | 4.5 | 24.3 | 5.4 | 0.02 |
|  | No. | % | No. | % | P |
| Cancer Site |  |  |  |  | 0.54 |
| Nasopharynx | 2 | 5 | 1 | 12.5 |  |
| Oropharynx | 24 | 60 | 2 | 25 |  |
| Glottis | 4 | 10 | 1 | 12.5 |  |
| CUP | 2 | 5 | 2 | 25 |  |
| Sinonasal | 2 | 5 | 1 | 12.5 |  |
| Skin | 2 | 5 | - | - |  |
| Salivary Gland | 1 | 2.5 | 1 | 12.5 |  |
| T stage |  |  |  |  | 0.57 |
| TX | 2 | 5 | - | - |  |
| T0-2 | 28 | 70 | 5 | 62.5 |  |
| T3-4 | 10 | 25 | 3 | 37.5 |  |
| N stage |  |  |  |  | 0.04 |
| N0-1 | 14 | 35 | 6 | 75 |  |
| N2-3 | 26 | 65 | 2 | 25 |  |
| Grade |  |  |  |  | 0.11 |
| Well differentiated | 4 | 10 | 0 | 0 |  |
| Mod differentiated | 12 | 30 | 2 | 25 |  |
| Poorly differentiated | 15 | 37.5 | 1 | 12.5 |  |
| Unknown | 9 | 22.5 | 5 | 62.5 |  |
| HPV-status |  |  |  |  | 0.53 |
| Positive | 23 | 57.5 | 3 | 37.5 |  |
| Negative | 2 | 5 | 1 | 12.5 |  |
| Not tested | 15 | 37.5 | 4 | 50 |  |
| ECOG PS |  |  |  |  | 0.18 |
| 0 | 25 | 62.5 | 6 | 75 |  |
| 1 | 13 | 32.5 | 1 | 12.5 |  |
| 2 | - | - | 1 | 12.5 |  |
| Unknown | 2 | 5 | - | - |  |
| SM depleted* |  |  |  |  | 0.28 |
| Yes | 12 | 30 | 4 | 50 |  |
| No | 28 | 70 | 4 | 50 |  |
| Induction chemotherapy |  |  |  |  | 0.22 |
| Yes | 4 | 10 | - | - |  |
| No | 36 | 90 | 8 | 100 |  |

### Whole body BIA vs. CT analysis

At the time of enrollment, no patients were underweight (BMI < 18.5 kg/m^2^), 8 patients (17%) had normal BMI (18.5-24.9), 23 patients (48%) were overweight (BMI 25-29.9), and 17 patients (35%) were obese (BMI ^3^ 30). By CT analysis, 16 patients (33%) were sarcopenic at the time of enrollment. Full body composition details measured by CT and BIA are shown in Table 2. Age, height, tumor site, T stage, N stage, grade, HPV status, smoking status, and use of induction chemotherapy did not vary by sarcopenic status (Supplementary Table 1).

**Table 2.** Pre-treatment Body Composition. Abbreviations: BMI, body mass index; CT, computed tomography; CSA, cross sectional area; SMI, skeletal muscle index; SM, skeletal muscle; ADI, adipose index; BIA, bioelectrical impedance analysis; FFM, fat free mass. Data represent mean ± SD. P-values calculated using Student’s t-test for continuous variables, Fisher’s Exact Test for categorical variables.

|  | Body Composition |  |  | P-value (sex) |
| --- | --- | --- | --- | --- |
|  | Total<br>N=48 | Male<br>N=40 | Female<br>N=8 |  |
| BMI categories, n (%) |  |  |  | 0.002 |
| <18.5 | 0 (0%) | 0 (0%) | 0 (0%) |  |
| 18.5-24.9 | 8 (17%) | 3 (7.5%) | 5 (62%) |  |
| 25-29.9 | 23 (48%) | 22 (55%) | 1 (13%) |  |
| ≥30 | 17 (35%) | 15 (37.5%) | 2 (25%) |  |
| CT measures, mean ± SD |  |  |  |  |
| L3 Muscle CSA (cm<sup>2</sup>) | 165.5 ± 41.0 | 178.8 ± 30.1 | 99.2 ± 13.3 | <.0001 |
| SMI (cm<sup>2</sup>/m<sup>2</sup>) | 53.0 ± 9.8 | 55.9 ± 7.7 | 38.5 ± 5.3 | <.0001 |
| Sarcopenia, n (%) | 16 (33%) | 12 (30%) | 4 (50%) | 0.41 |
| L3 Fat CSA (cm<sup>2</sup>) | 345.7 ± 135.4 | 353.8 ± 135.2 | 305.0 ± 137.9 | 0.38 |
| ADI (cm<sup>2</sup>/m<sup>2</sup>) | 112.6 ± 45.2 | 111.1 ± 43.1 | 120.2 ± 57.5 | 0.68 |
| BIA measures, mean ± SD |  |  |  |  |
| SM mass (kg) | 29.1 ± 7.3 | 31.6 ± 5.1 | 16.9 ± 2.7 | <.0001 |
| SM mass (% body weight) | 14.9 ± 2.2% | 15.6 ± 1.4% | 11.6 ± 2.4% | 0.002 |
| SM mass index (kg/m<sup>2</sup>) | 9.3 ± 1.8 | 9.9 ± 1.3 | 6.5 ± 0.9 | <.0001 |
| Fat free mass (kg) | 60.6 ± 12.4 | 64.9 ± 8.3 | 39.2 ± 4.1 | <.0001 |
| Fat free mass (% body weight) | 67.1 ± 6.6% | 67.9 ± 5.7% | 63.1 ± 9.3% | 0.2 |
| FFM index (kg/m<sup>2</sup>) | 19.3 ± 2.7 | 20.2 ± 1.9 | 15.0 ± 2.0 | <.0001 |
| Fat mass (kg) | 30.4 ± 10.5 | 31.7 ± 10.3 | 24.1 ± 9.8 | 0.07 |
| Fat mass (% body weight) | 32.9 ± 6.6% | 32.1 ± 5.7% | 36.9 ± 9.3% | 0.2 |
| Fat mass index (kg/m<sup>2</sup>) | 9.8 ± 3.3 | 9.8 ± 3.1 | 9.3 ± 4.1 | 0.74 |

To determine whether BIA could accurately replicate CT-derived body composition metrics, we conducted a series of linear regressions. We first evaluated the relationships between BIA measures of SM mass, FFM, and FM against unnormalized CT-derived SM and adipose CSA (Figure 1). BIA SM mass was highly correlated to CT SM CSA with SM mass (kg) = 0.511 + 0.173 x [SM CSA at L3 (cm^2^)], *r* = 0.97; *p* < 0.0001. BIA FFM was also closely associated with CT SM CSA; FFM (kg) = 12.177 + 0.293 x [SM CSA at L3 (cm^2^)], *r* = 0.97; *p* < 0.0001. BIA accurately represented CT measures of fat mass; FM mass (kg) = 4.882 + 0.074 x [Adipose CSA at L3 (cm^2^)], *r* = 0.95; *p* < 0.0001.

**Figure 1.**
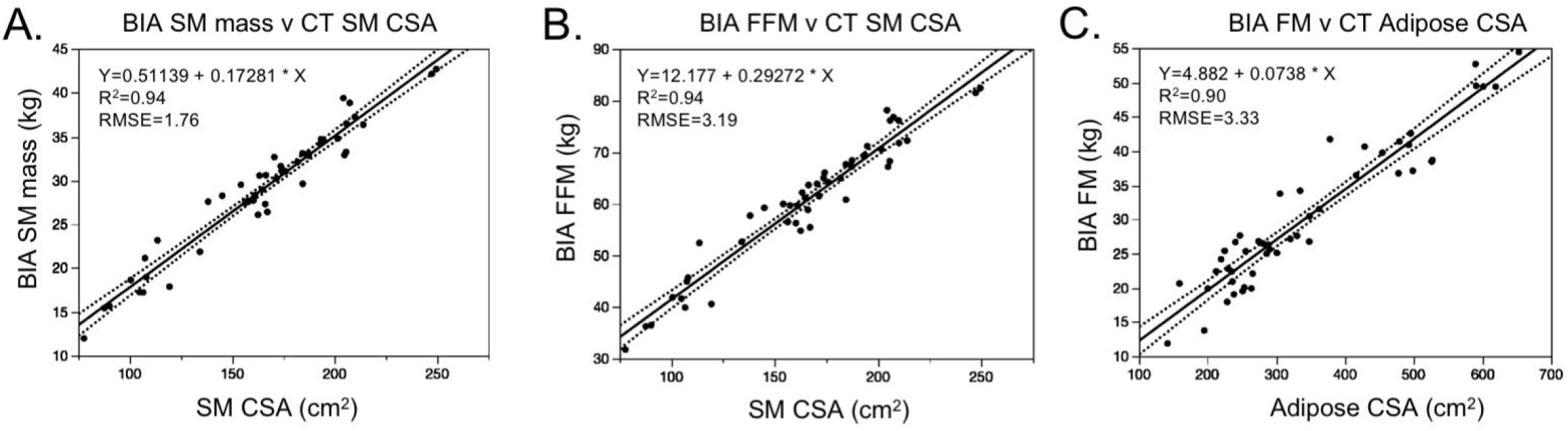
BIA measures of body composition compared to CT-based estimates and BMI. **A**, Skeletal Muscle (SM), **B**, Fat Free Mass (FFM) and **C**, Fat Mass (FM) measured by BIA compared to SM and FM estimates derived from lumbar CT scans. **D**, SM, **E**, FFM, and **F**, FM measured by BIA compared to BMI (n=48).

Because thresholds for survival-associated sarcopenia are based on normalized body composition indices, we repeated these correlations using normalized tissue compartment measures. For BIA, this includes the SM index (SMI), FFM index (FFMI), and FM index (FMI), each of which is automatically calculated by dividing the measured mass value by the square of height in meters. Linear regression revealed close associations between BIA and CT (Figure 2A-2C):

**Figure 2.**
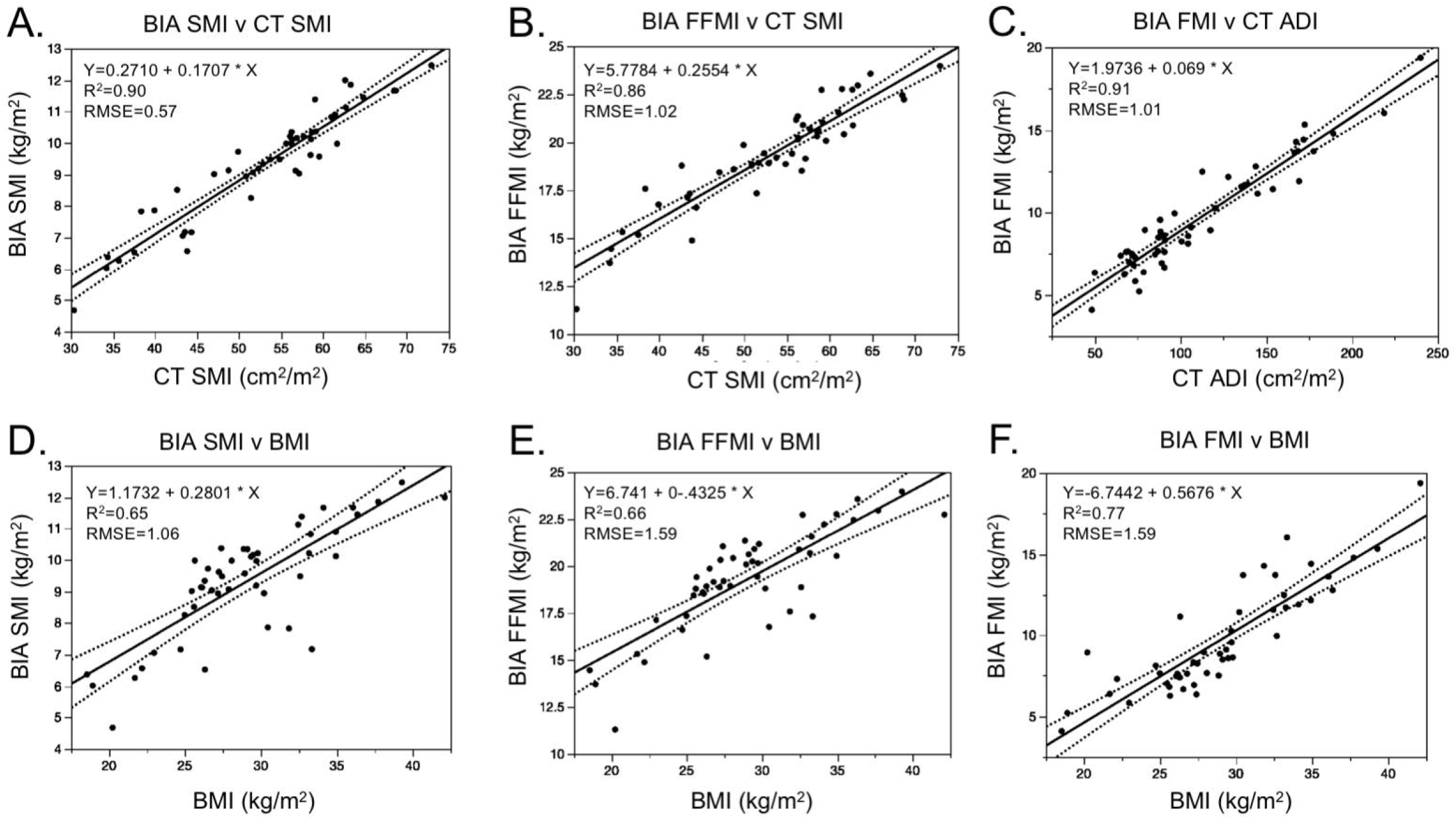
Normalized BIA measures of body composition compared to CT-based estimates and BMI. **A**, Skeletal Muscle Index (SMI), **B**, Fat Free Mass Index (FFMI) and **C**, Fat Mass Index (FMI) measured by BIA compared to SMI and adipose index (ADI) estimates derived from lumbar CT scans. **D**, SMI, **E**, FFMI, and **F**, FMI measured by BIA compared to BMI (n=48).

- BIA SMI (kg/m^2^) = 0.271 + 0.171 x [CT SMI (kg/m^2^)]; *r* = 0.95, *p* < 0.0001
- BIA FFMI (kg/m^2^) = 5.778 + 0.255 x [CT SMI (kg/m^2^)]; *r* = 0.93, *p* < 0.0001
- BIA FMI (kg/m^2^) = 1.974 + 0.069 x [CT ADI (kg/m^2^)]; *r* = 0.95, *p* < 0.0001

We then tested the association between BIA measures of body composition and BMI, to identify which tissue compartments are best estimated by BMI. BIA measures were less closely associated with BMI than CT, with the strongest association seen between BIA FMI and BMI (Figure 2D-2F):

- BIA SMI (kg/m^2^) = 1.173 + 0.128 x [BMI (kg/m^2^)]; *r* = 0.80, *p* < 0.0001
- BIA FFMI (kg/m^2^) = 6.741 + 0.433 x [BMI (kg/m^2^)]; *r* = 0.81, *p* < 0.0001
- BIA FMI (kg/m^2^) = 6.744 + 0.568 x [BMI (kg/m^2^)]; *r* = 0.88, *p* < 0.0001

### Segmental BIA vs. CT analysis

The 8-electrode design of the BIA device used in this study allows for independent estimates of SM mass and FM in each limb and the torso. To determine which body segment provided the best reflection of CT-based SM mass, we ran a series of linear regressions, plotting SM mass from the torso, right and left legs, and right and left arms against CT SM CSA at the L3 level. These regressions show that each body segment is independently closely associated with L3 SM CSA. The left and right arms have the highest correlation (left arm, R^2^ = 0.92, RMSE = 0.17; right arm, R^2^ = 0.92, RMSE = 0.16), whereas the legs exhibit the lowest (left leg, R^2^ = 0.90, RMSE = 0.41; right leg, R^2^ = 0.88, RMSE = 0.46). The torso demonstrated the highest error among segments (R^2^ = 0.91, RMSE = 1.14), although this is likely driven by having the largest absolute mass value.

### BIA-defined sarcopenia

Having observed close correlation between BIA and CT measured SM mass estimates, we then sought to use these relationships to define sex-specific BIA cutoffs for sarcopenia. For each BIA measure of SM mass or FFM, we performed ROC analysis using CT-defined sarcopenia as the “positive” level. The maximum Youden Index value was used to select the optimum cut-off defining sarcopenia. Sensitivity and specificity were then calculated for each regressor using the optimized threshold. For men, optimal cutoffs for SM mass and FFM, both raw and normalized to the square of height in meters, demonstrated high sensitivity and specificity for defining sarcopenia, as evidenced by ROC c-statistics > 0.92 (Table 3). The best discriminator for sarcopenia, as defined by the highest ROC c-statistic, was SM mass index < 9.19 kg/m^2^ for men (c-statistic 0.964, sensitivity 91.7%, specificity 92.9%). Analysis of the optimized cut-off in women was limited by the small number of women in the study. Despite this, discriminatory performance was also highest when using SM mass index < 6.53 kg/m^2^ (c-statistic 1.0, sensitivity 100%, specificity 0%).

**Table 3.**
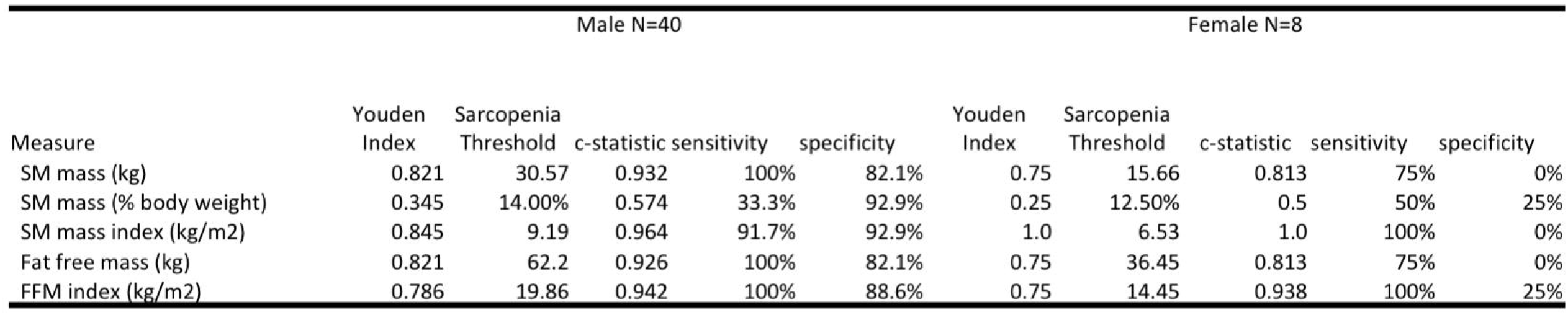
BIA measures of SM depletion. Receiver operator curve analysis to identify optimal sex-specific BIA thresholds for SM depletion. Sensitivity and specificity reflect values at the maximum Youden Index for each regressor. Abbreviations: SM, skeletal muscle; FFM, fat free mass.

### Muscle, Fat, and Weight Loss during RT

Patients lost an average of 5.7 ± 5.8 kg during treatment, with mean BMI dropping from 29.4 ± 5.5 kg/m^2^, to 27.8 ± 4.7 kg/m^2^ at treatment completion (Supplementary Figure 1). SM mass decreased by a mean of 1.5 ± 1.9 kg during treatment from 29.8 ± 7.6 kg to 28.3 ± 6.7 kg. FFM decreased by a mean of 2.6 ± 3.3 kg from a mean of 61.8 ± 13.0 kg to 59.2 ± 11.6 kg. Mean FM loss was 2.2 ± 2.6 kg, from an initial mean FM of 30.7 ± 12.0 kg to 28.5 ± 10.9 kg. Patients lost an average of 0.8 ± 1.8 kg of total body water during RT. Using the BIA measured sex-specific SMI cutoffs for sarcopenia described above, the number of patients with sarcopenia increased from 16 (34.7%) prior to RT, to 23 (50%) post-RT (n=46). Eight patients developed sarcopenia during RT, whereas one sarcopenic patient no longer met the criteria for sarcopenia at treatment completion. Therapeutic feeding tubes were placed in 7 non-sarcopenic patients (23.3%) and 3 sarcopenic patients (18.8%).

## Discussion

To our knowledge this study is the first prospective validation of BIA as a measure of sarcopenia. BIA measures of body composition were very strongly correlated with CT-based estimates, validating BIA as an accurate alternative measure for sarcopenia in a clinical head and neck cancer population. The close associations between BIA and CT were observed despite methodology that did not control for variations in eating, drinking, or exercising prior to BIA. Given the difficulties in maintaining nutrition exhibited by many head and neck cancer patients, we felt it is important to derive meaningful results without the need for fasting or other metabolic challenges. BIA was easy to perform and was readily integrated into a busy radiation oncology clinic; in our study weekly BIA was performed by registered dieticians and took approximately 2 minutes per visit. In contrast to DEXA, which is seldom available in radiation centers, or CT or MRI, which are expensive to perform and require dedicated post-hoc analysis, BIA provided rapid results that were immediately interpretable. Given the plurality of studies reporting an association between depleted SM mass and shortened survival, interrogation of body composition can help clinicians with *a priori* risk stratification and may prove useful in selecting optimal treatment regimens amid ongoing efforts to de-escalate therapy.

Once we had validated BIA measures of SM mass, we then used the relationship between BIA and CT to identify normalized BIA-derived thresholds for identifying sarcopenia. The thresholds identified in this study fall within the range of “moderate sarcopenia”, identified by Janssen and colleagues as being associated with physical disability among older adults.^17^ That normalized SM mass values in this range are associated with poor outcomes in both populations supports the interpretation that low muscle mass is a common indicator of underlying pathology. However, because head and neck cancer is increasingly common among younger patients, both age- and cancer-specific thresholds for sarcopenia may be necessary for clinical utility.

Our results show no association between sarcopenia and other factors prognostic for survival, including HPV status, smoking history, age, and tumor stage. Sarcopenia was equally prevalent among patients with HPV-associated cancers, which often present with a small primary tumor and involved neck lymph nodes. Whereas our retrospective data suggested that sarcopenia portends worse outcomes in patients with HPV-associated cancers ^1^, recently published retrospective data question this association.^4^ Thus, the prognostic value of sarcopenia among HPV-positive cancers, which are typically associated with improved survival, remains uncertain and should be the subject of future prospective analysis.

As a secondary outcome, we evaluated whether BIA was sensitive enough to monitor individual changes in body composition across treatment. At present, body weight is the primary indicator used to identify patients who need additional nutrition support, including dietetic intervention or feeding tube placement. However, our prior retrospective report found that increased weight loss was associated with improved survival and cancer control.^1^ This is because the greatest weight loss was observed in obese patients, and obesity itself was prognostic for better cancer-free and overall survival. In contrast, those patients who developed sarcopenia between starting RT and their follow up imaging had poorer survival, reinforcing the importance of knowing which tissues are driving weight loss. BIA offers more information than body weight and may identify patients approaching the sarcopenia threshold. This could potentially help stratify patients in nutritional and supportive care trials investigating nutritional, exercise-based, and pharmacologic approaches to improving muscle mass. It seems logical that strength training and adequate protein intake may help, and the DAHANCA group is investigating this ^18^.

### Limitations

The low number of women participants precludes generalizability of our findings to female patients with head and neck cancer. Specific thresholds for BIA-determined sarcopenia need to be validated and refined in larger studies that include greater numbers of women patients.

Evaluation of this technology in other cancer sites may demonstrate the need for cancer-specific and age-specific sarcopenia cut-offs. Our study also did not include post-RT imaging to validate the accuracy of post-treatment changes in body composition or the ability of BIA to identify new-onset sarcopenia. Finally, this study did not include the long term oncologic and survival outcomes for this population. Thus, we could not validate the previously described relationship between sarcopenia and outcome in this population.

## Conclusions

In this prospective validation study, BIA was closely correlated with CT-derived measures of SM mass and FM. BIA demonstrated high sensitivity and specificity for identifying patients with sarcopenia, a negative prognostic factor in head and neck cancer. The use of BIA may be a practical solution for identifying patients with sarcopenia in routine clinical practice.

## Data Availability

The primary data that support the findings of this study are available on request from the corresponding author.

## Acknowledgements

Study supported by funding from the National Institute for Dental and Craniofacial Research Award (1R01DE025248-01/R56DE025248) and Academic-Industrial Partnership Award (1R01 DE028290-01), the National Science Foundation (NSF), Division of Mathematical Sciences, Joint NIH/NSF Initiative on Quantitative Approaches to Biomedical Big Data (QuBBD) Grant (NSF 1557679), the NIH Big Data to Knowledge (BD2K) Program of the National Cancer Institute (NCI) Early Stage Development of Technologies in Biomedical Computing, Informatics, and Big Data Science Award (1R01CA214825), the NCI Early Phase Clinical Trials in Imaging and Image-Guided Interventions Program (1R01CA218148), the NIH/NCI Cancer Center Support Grant (CCSG) Pilot Research Program Award from the UT MD Anderson CCSG Radiation Oncology and Cancer Imaging Program (P30CA016672), the NIH/NCI Head and Neck Specialized Programs of Research Excellence (SPORE) Developmental Research Program Award (P50 CA097007) and the National Institute of Biomedical Imaging and Bioengineering (NIBIB) Research Education Program (R25EB025787).

